# Alterations in Resting-State Functional Connectivity and Dynamics in Schizophrenia as a Result of Global not Local Processes

**DOI:** 10.1101/2023.12.08.23299714

**Authors:** Christoph Metzner, Cristiana Dimulescu, Fabian Kamp, Sophie Fromm, Peter J Uhlhaas, Klaus Obermayer

**Affiliations:** Neural Information Processing Group, Institute of Software Engineering and Theoretical Computer Science, Technische Universität Berlin, Berlin, Germany; Department of Child and Adolescent Psychiatry, Charité – Universitätsmedizin Berlin, corporate member of Freie Universität Berlin and Humboldt-Universität zu Berlin, Berlin, Germany; School of Physics, Engineering and Computer Science, University of Hertfordshire, Hatfield, United Kingdom; Bernstein Center for Computational Neuroscience Berlin, Berlin, Germany; Max Planck School of Cognition, Max Planck Institute for Human Cognitive and Brain Science, Leipzig, Germany; Center for Lifespan Psychology, Max Planck Institute for Human Development, Berlin, Germany; Department of Psychiatry and Psychotherapy, Charité – Universitätsmedizin Berlin, corporate member of Freie Universität Berlin and Humboldt-Universität zu Berlin, Berlin, Germany; Institute of Neuroscience and Psychology, University of Glasgow, Glasgow, United Kingdom

**Keywords:** schizophrenia, resting-state fMRI, computational model, large-scale networks, functional connectivity, temporal dynamics

## Abstract

We examined changes in large-scale functional connectivity and temporal dynamics and their underlying mechanisms in schizophrenia (ScZ) through measurements of resting-state functional magnetic resonance imaging (rs-fMRI) data and computational modelling. The rs-fMRI measurements from patients with chronic ScZ (n=38) and matched healthy controls (n=43), were obtained through the public schizConnect repository. Computational models were constructed based on diffusion-weighted MRI scans and fit to the experimental rs-fMRI data. We found decreased large-scale functional connectivity across sensory and association areas and for all functional subnetworks for the ScZ group. Additionally global synchrony was reduced in patients while metastability was unaltered. Perturbations of the computational model revealed that decreased global coupling and increased background noise levels both explained the experimentally found deficits better than local changes to the GABAergic or glutamatergic system. The current study suggests that large-scale alterations in ScZ are more likely the result of global rather than local network changes.

## 1 INTRODUCTION

ScZ is a severe mental disorder with a high burden of disease (Lopez and Murray (1998); Charlson et al. (2018)). However, the underlying mechanisms remain elusive. While no single brain area accounting for the heterogeneous symptom profiles has been identified, the notion that ScZ can be understood in terms of a general dysconnectivity has emerged (Friston et al. (1995); Friston (1999); Bullmore et al. (1997); Pettersson-Yeo et al. (2011)).

Experimental evidence for the dysconnection hypothesis comes from neuroimaging studies. Analyses of resting-state fMRI connectivity have shown widespread changes of functional connectivity. However, there is still a debate whether correlations of neural activity between regions are decreased (Liang et al. (2006); Bluhm et al. (2007)) or increased in ScZ (Zhou et al. (2007)). There is also growing evidence for possible longitudinal changes of functional connectivity over the course of the disorder. Anticevic et al. (2015) demonstrated that prefrontal cortical connectivity is increased in early-course ScZ while the opposite pattern was observed in chronic ScZ patients. Going beyond pairwise correlations between brain regions, graph theoretic measurements have identified reductions in integration, hierarchy, clustering, efficiency and small-worldness (Bassett et al. (2008); Liu et al. (2008); Bullmore and Sporns (2009); Lynall et al. (2010)).

Yet, the origin of functional dysconnectivity patterns in ScZ is still unclear. One hypothesis is that cellular and synaptic changes associated with ScZ disrupt local processing and thus impact on large-scale connectivity. Indeed changes at the microcircuit level have been identified in ScZ. Excitatory and inhibitory neurotransmission is disturbed, for example a reduced excitatory drive onto GABAergic inhibitory neurons (Chung et al. (2016, 2022)) and a decreased inhibitory output (Hashimoto et al. (2003); Morris et al. (2008); Moyer et al. (2012)). Changes to the glutamatergic system, such as increased recurrent excitation, have been suggested to lead to deficits in large-scale connectivity with a gradient along the cortical hierarchy (Yang et al. (2016)).

Computational models of large-scale brain circuits can be used to investigate dynamical circuit mechanisms linking local ScZ-associated alterations to global changes in the functional organisation of the brain. Leveraging such computational models, studies have shown that decreases in global inter-regional connectivity strengths can lead to wide-spread functional disruptions (Cabral et al. (2013)), increased global signal variance (Yang et al. (2014)) and altered topological characteristics of functional brain networks (Cabral et al. (2012b,a)) resembling ScZ. However, except for Yang et al. (2014), these studies only investigated a global scaling of the inter-regional connectivity. Yang et al. (2014) manipulated local and global neuronal coupling and demonstrated that both could increase signal variance as seen in ScZ but did not explore their potentially differential effects on large-scale functional connectivity. Thus, so far the effect of ScZ-associated local changes to glutamatergic and GABAergic neurotransmission and the effect of increased background noise on large-scale functional connectivity has not been explored.

To address this question, we quantified functional connectivity differences in a data set of healthy controls and chronic ScZ patients. We then implemented local microcircuit and global network parameter changes in a computational model of large-scale cortical dynamics and compare the resulting connectivity changes to the experimental data. Furthermore, we also explored the temporal dynamics of the resting-state brain and characterised potential deficits in large-scale synchrony and metastability in ScZ patients and compared them to the different computational models, thus identifying mechanistic links underlying these changes.

## 2 MATERIAL AND METHODS

### 2.1 Patient Sample

The study sample was collected through the Center for Biomedical Research Excellence (COBRE) led by Dr. Vince Calhoun (more information here: *http://fcon1000.projects.nitrc.org/indi/retro/cobre.html*) and obtained from the SchizConnect database (*http://schizconnect.org*). This sample has previously been used by our group to explore structural deficits in patients with ScZ (Dimulescu et al. (2021)). From the sample of 43 patients and 43 healthy control participants, we excluded 5 patients due to missing resting-state functional MRI (rs-fMRI) data or artefacts/excessive motion identified during the pre-processing. We thus analyzed a final sample of 43 healthy control subjects and 38 patients with schizophrenia, which we will refer to as the COBRE sample. All patients were receiving antipsychotic medication (see Table 1). Symptom severity in patients was assessed using the Positive and Negative Syndrome Scale (PANSS) (Kay et al. (1989)). Written informed consent was obtained from all participants, and the study was reviewed and approved by the Institutional Review Board of the University of New Mexico.

**Table 1.** Demographics and clinical characteristics. Data are shown as mean(standard deviation). Age differences between groups were compared using an independent samples t-test and differences in gender distribution using a chi-square test. Antipsychotic medication is reported as chlorpromazine (CPZ)-equivalent dosage.

|  | HC | ScZ | Statistics, *p* value |
| --- | --- | --- | --- |
| Group size | 43 | 38 | - |
| Age (y) | 36.70(11.04) | 38.97(13.67) | *t*=0.82, *p*=0.41 |
| Gender | 11F/32M | 10F/27M | $\chi^2=0.02$, *p*=0.88 |
| PANSS positive | - | 14.92(5.04) | - |
| PANSS negative | - | 14.81(5.31) | - |
| PANSS general | - | 29.49(8.37) | - |
| PANSS total | - | 59.22(78) | - |
| CPZ-equivalent dosage | - | 396.26(330.91) | - |
| Illness duration (y) | - | 17.19(12.61) | - |

### 2.2 Anatomical data

Data collection for the COBRE sample was performed using a Siemens Magnetom Trio 3T MR scanner. Structural images (high resolution T1-weighted) were acquired using a five-echo MPRAGE sequence with the following parameters: repetition time (TR) = 2530 ms; echo time (TE) = 1.64, 3.5, 5.36, 7.22, 9.08 ms; inversion time (TI) = 1200 ms; flip angle (FA) = 7^*?*^; field of view (FOV) = 256 mm × 256 mm; matrix = 256 × 256; slice thickness = 1 mm; 192 sagittal slices. Diffusion tensor imaging (DTI) data were acquired using a single-shot EPI sequence with TR/TE = 9000/84 ms; FA = 90^*?*^; FOV =256 mm × 256 mm; matrix = 128 × 128; slice thickness = 2 mm without gap; 72 axial slices; 30 non-collinear diffusion gradients (b = 800 s/mm^2^) and 5 non-diffusion-weighted images (b = 0 s/mm^2^) equally interspersed between the 30 gradient directions. For more information see also Cetin et al. (2014).

For model validation we additionally used a subset of 156 healthy participants from the human connectome project (HCP), which we will refer to as the HCP sample. The diffusion-weighted data were collected with multiband diffusion sequence (HCP version available at *http://www.cmrr.umn.edu/multiband*). Three different gradient tables are used, each with 90 diffusion weighting directions and six *b* = 0 acquisitions. More information can be found at *https://www.humanconnectome.org/study/hcp-young-adult/document/1200-subjects-data-release*.

### 2.3 Resting-state functional MRI data

COBRE data was acquired using single-shot full k-space echo-planar imaging (EPI) with ramp sampling correction using the intercomissural line (AC-PC) as a reference (TR: 2 s, TE: 29 ms, matrix size: 64x64, 32 slices, voxel size: 3x3x4 mm^3^). The resting-state scans were acquired in the axial plane with with an ascending slice order (multi slice method; interleaved). For more information see Aine et al. (2017). For the COBRE data set, we preprocessed the rsfMRI data using the FSL FEAT toolbox (Woolrich et al. (2001)). For each data set, we discarded the first five volumes. We corrected head motion using the FSL McFLIRT algorithm and subsequently high-pass filtered the data with a filter cutoff of 100 s. We linearly registered each functional image to the corresponding anatomical image of that subject using FLIRT. We then used the mean volume of the data to create a brain mask using BET. Using the ICA FIX FSL toolbox (Griffanti et al. (2014); Salimi-Khorshidi et al. (2014)), we conducted MELODIC ICA and removed artefactual components (motion, non-neuronal physiological artefacts, scanner artefacts, and other nuisance sources). Finally, we transformed the high-resolution mask volumes from MNI to individual subject functional space and extracted the average BOLD time courses for each cortical region in the AAL2 parcellation scheme using the fslmeants command from Fslutils.

Acquisition details for the functional MRI data from the HCP S1200 release can be found here: *https://www.humanconnectome.org/study/hcp-young-adult/document/1200-subjects-data-release*. For the HCP data set, we used the data preprocessed according to Glasser et al. (2013) and extracted the average BOLD time courses for each cortical region as described above.

### 2.4 Measures of connectivity and temporal dynamics

We used the average global brain connectivity (GBC) measure (Cole et al. (2010, 2011)) to assess the changes in connectivity strength. To assess alterations in temporal dynamics we used synchrony and metastability (Deco et al. (2017)). Because of the computational model being restricted to cortical areas, we also restricted our connectivity analysis to cortical areas. However, including subcortical regions did not substantially change the findings (see Supplementary Material).

Specifically, we define the functional connectivity matrix (FC) as the matrix of Pearson correlations of the BOLD signal between two brain areas over the whole time range of acquisition. From the FC matrices we calculate the global brain connectivity (GBC) of a single brain region *i* as follows (see also Cole et al. (2010, 2011)):

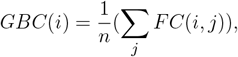

where *n* is the number of regions. The average global GBC can then be defined as the average GBC over all cortical regions *i*. To calculate the average GBC for a functional subnetwork or generally a set of regions, one simply averages over the regional GBC values for the respective regions.

To assess the temporal dynamics of the functional networks, we used the Kuramoto order parameter as a measure of synchrony and its standard deviation as a measure of metastability, i.e. the variability of the states of phase configurations over time (see for example Deco et al. (2017)). Here the Kuramoto order parameter *R*(*t*) is defined as

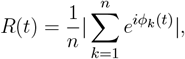

where again *n* is the number of regions and *ϕ*_*k*_(*t*) is the instantaneous phase of the BOLD signal in region *k*. It measures the global level of synchronization of the BOLD signals from all regions, where a low level close to 0 reflects an almost uniform distribution of the signal phases, and a high value close to 1 reflects near equality of the signal phases. To calculate *R*, we band-pass filtered the signal in the narrowband 0.04-0.07 Hz (see Deco et al. (2017)) and then extracted the instantaneous phases of the signals at every time step using the Hilbert transform.

### 2.5 Computational network model

We use a whole-brain network model, where the connectivity, connection strength and delay between network nodes (i.e. brain regions) is derived from brain imaging data. As a model of single-node activity dynamics we employ a mean-field description of a spiking neural network of an excitatory and an inhibitory neural population, where the individual neurons are described by the adaptive exponential integrate-and-fire model (AdEx model; Brette and Gerstner (2005)), developed in our group (Augustin et al. (2017); Cakan and Obermayer (2020)). The following section describes the model in detail.

#### 2.5.1 Single-Node model

A mean-field neural mass model based on a spiking network of coupled excitatory and inhibitory populations, the so-called ALN model (Augustin et al. (2017)), was implemented. The mean-field description offers a drastic speed-up of simulations on the order of about 4 orders of magnitude compared to the spiking model while still retaining its dynamical states and its biophysical parameters. The model has been extensively validated against simulations with the detailed spiking network and overall shows very good agreement (Cakan and Obermayer (2020)).

The mean-field reduction of the spiking neural network utilises the Fokker-Planck approach, i.e. the fact that in the limit of an infinite network size and under the assumption of a sparse, random connectivity, the distribution *p*(*V*) of the membrane potentials and the mean firing rate *r*_*a*_ of a population *a*, can be described by a Fokker-Planck equation (Brunel (2000)). However, to calculate the potential distribution a partial differential equation has to be solved, which is computationally costly. Therefore, the dynamics of a population is captured by a low-dimensional linear-nonlinear cascade model, and can be described by a set of ordinary differential equations (Fourcaud-Trocmé et al. (2003); Ostojic and Brunel (2011)). The mathematical derivation and the underlying assumptions have been detailed in (Augustin et al. (2017)), and we will only provide the final set of model equations in this manuscript.

A single network node in the whole-brain model is represented by the population activity of two interconnected neural populations, an excitatory population *E* and an inhibitory population *I*. The dynamics of the membrane currents of a population *a* ∈ {*E, I*}, are governed by the following equations:

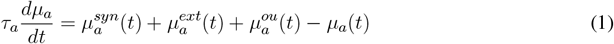

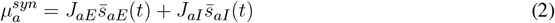

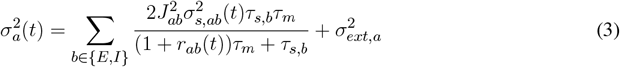

In the above equations *μ*_*a*_ describes the total mean membrane currents, 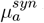 the currents from synaptic activity, 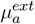 the currents from any sources of external input, 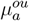 the external noise input, *τ*_*m*_ the membrane time constant (calculated from the membrane capacitance *C* and the leak conductance *g*_*L*_), and *τ*_*s,b*_ the synaptic time constant. Furthermore, 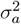 is the variance of the membrane currents, and *J*_*ab*_ represent the maximum synaptic current when all synapses from population b to population a are active. The dynamics of the synapses are described by

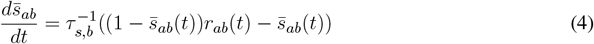

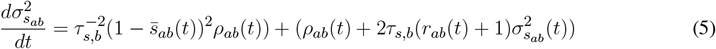

where 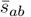 represents the mean of the fraction of all active synapses, which lies in the range [0, 1], with the extreme cases being no active synapses and no inactive synapses, respectively. Furthermore, 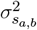 is the variance of *s*_*ab*_.

The timescale *τ*_*a*_ = Φ_*T*_ (*μ*_*a*_, *σ*_*a*_) of the input-dependent adaptation, the average membrane potential 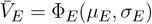, and the instantaneous population spike rate *r*_*a*_ = Φ_*r*_(*μ*_*a*_, *σ*_*a*_) are computed every time step by means of precomputed transfer functions. The mean *r*_*ab*_ and the variance *ρ*_*ab*_ of the effective input rate from population b to population a can be described by

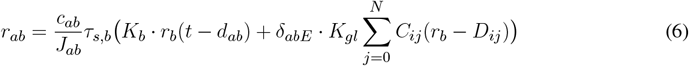

and

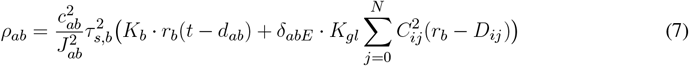

given a certain delay for the spike transmission *d*_*ab*_. Here *c*_*ab*_ represent the amplitude of the post-synaptic current resulting from one individual spike (for *s*_*ab*_ = 0). Furthermore, *K*_*gl*_ scales the global coupling in the network, and *C*_*ij*_ and *D*_*ij*_ define the connection strengths and the connection delays between regions, estimated from the fibre count and fibre length matrices, respectively. Finally, *δ*_*abE*_ = 1 for *a* = *b* = *E* and 0 otherwise restricting coupling between regions to be exclusively from excitatory to excitatory populations.

The adaptive exponential integrate-and-fire model explicitly accounts for the evolution of a slow adaptation currents that represents both subthreshold and spike-triggered adaptation currents. The subthreshold adaptation current is described by the adaptation conductance *α* and the spike-triggered adaptation current is denoted by *β*. In the limit of infinite population sizes, an adiabatic approximation can be employed to describe the mean adaptation current in terms of the mean population firing rate. The mean adaptation current *Ī*_*A*_ can be understood as an inhibitory membrane current whose dynamics are governed by

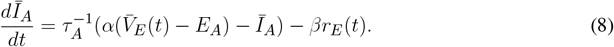

The individual populations *a* of a single region of the whole-brain network receive an external input current with a given mean 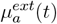 and a standard deviation 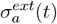. This background input current can be thought to represent baseline input from extracortical areas in the brain. Additionally, the regions also receive a noise input current 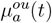 modelled as an Ornstein-Uhlenbeck process with a mean of 0 described by

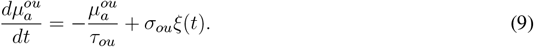

Here *ξ*(*t*) is a white noise process drawn from a normal distribution with a mean of 0 and a variance of 1. *σ*_*ou*_ determines the fluctuation amplitude of the noise around its mean.

To determine the mean external input to the E (*μ*_*Eext*_) and I (*μ*_*Iext*_) populations, the noise strength *σ*_*ou*_, the subthreshold adaptation conductance *α* and spike-triggered adaptation increment *β* parameters for the model in the control condition, we used an evolutionary optimization procedure as described in Cakan et al. (2022). We compared the simulated BOLD FC to the empirical rsfMRI data. We initialized the algorithm with a random population of *N*_*init*_ = 160 individuals and repeated the evolutionary block with *N*_*pop*_ = 80 individuals for 100 generations. Initial parameter values were selected from a uniform distribution across the following intervals for the model parameters: *μ*_*Eext*_ ∈ [0.0, 4.0] *mV/ms, μ*_*Iext*_ ∈ [0.0, 4.0] *mV/ms, σ*_*ou*_ ∈ [0.0, 0.3], *a* ∈ [0.0, 40.0] *nS*, and *b* ∈ [0.0, 40.0] *pA*. The global coupling strength was set as in Figure 2 of Cakan et al. (2022). All other model parameters were set as given in Table 1 in Cakan et al. (2022) and they are summarised in Table 2.

**Table 2.** Network parameters. Overview of the different parameter values for the whole-brain model employed here.

| Parameter | Value | Description |
| --- | --- | --- |
| $\mu_E^{ext}$ | 1.63 mV/ms | Mean external input to E |
| $\mu_I^{ext}$ | 0.05 mV/ms | Mean external input to I |
| $\sigma_{ou}$ | 0.19 | Noise strength |
| $\tau_{ou}$ | 5.0 ms | Noise time constant |
| $K_e$ | 800 | Number of excitatory inputs per neuron |
| $K_i$ | 200 | Number of inhibitory inputs per neuron |
| $C_{EE}, C_{IE}$ | 0.3 mV/ms | Maximum AMPA PSC amplitude |
| $C_{EI}, C_{II}$ | 0.5 mV/ms | Maximum GABA PSC amplitude |
| $J_{EE}$ | 2.4 mV/ms | Maximum synaptic current from E to E |
| $J_{IE}$ | 2.6 mV/ms | Maximum synaptic current from E to I |
| $J_{EI}$ | -3.3 mV/ms | Maximum synaptic current from I to E |
| $J_{II}$ | -1.6 mV/ms | Maximum synaptic current from I to I |
| $\tau_{s,E}$ | 2 ms | Excitatory synaptic time constant |
| $\tau_{s,I}$ | 5 ms | Inhibitory synaptic time constant |
| $d_E$ | 4 ms | Synaptic delay to excitatory neurons |
| $d_I$ | 2 ms | Synaptic delay to inhibitory neurons |
| $C$ | 200 pF | Membrane capacitance |
| $g_L$ | 10 nS | Leak conductance |
| $\tau_m$ | $C/g_L$ | Membrane time constant |
| $E_L$ | -65 mV | Leak reversal potential of the AdEx model |
| $\Delta_T$ | 1.5 mV | Threshold slope factor of the AdEx model |
| $V_T$ | -50 mV | Threshold voltage of the AdEx model |
| $V_s$ | -40 mV | Spike voltage threshold of the AdEx model |
| $T_{net}$ | 1.5 ms | Refractory time of the AdEx model |
| $\sigma^{ext}$ | 1.5 mV/$\sqrt{ms}$ | Standard deviation of external input |
| $E_A$ | -80 mV | Adaptation reversal potential |
| $\alpha$ | 28.26 nS | Subthreshold adaptation conductance |
| $\beta$ | 24.04 pA | Spike-triggered adaptation increment |
| $\tau_A$ | 200 ms | Adaptation time constant |
| $K_{gl}$ | 250.0 | Global coupling strength |
| $v_{gl}$ | 20.0 m/s | Global signal speed |

**Figure 1.**
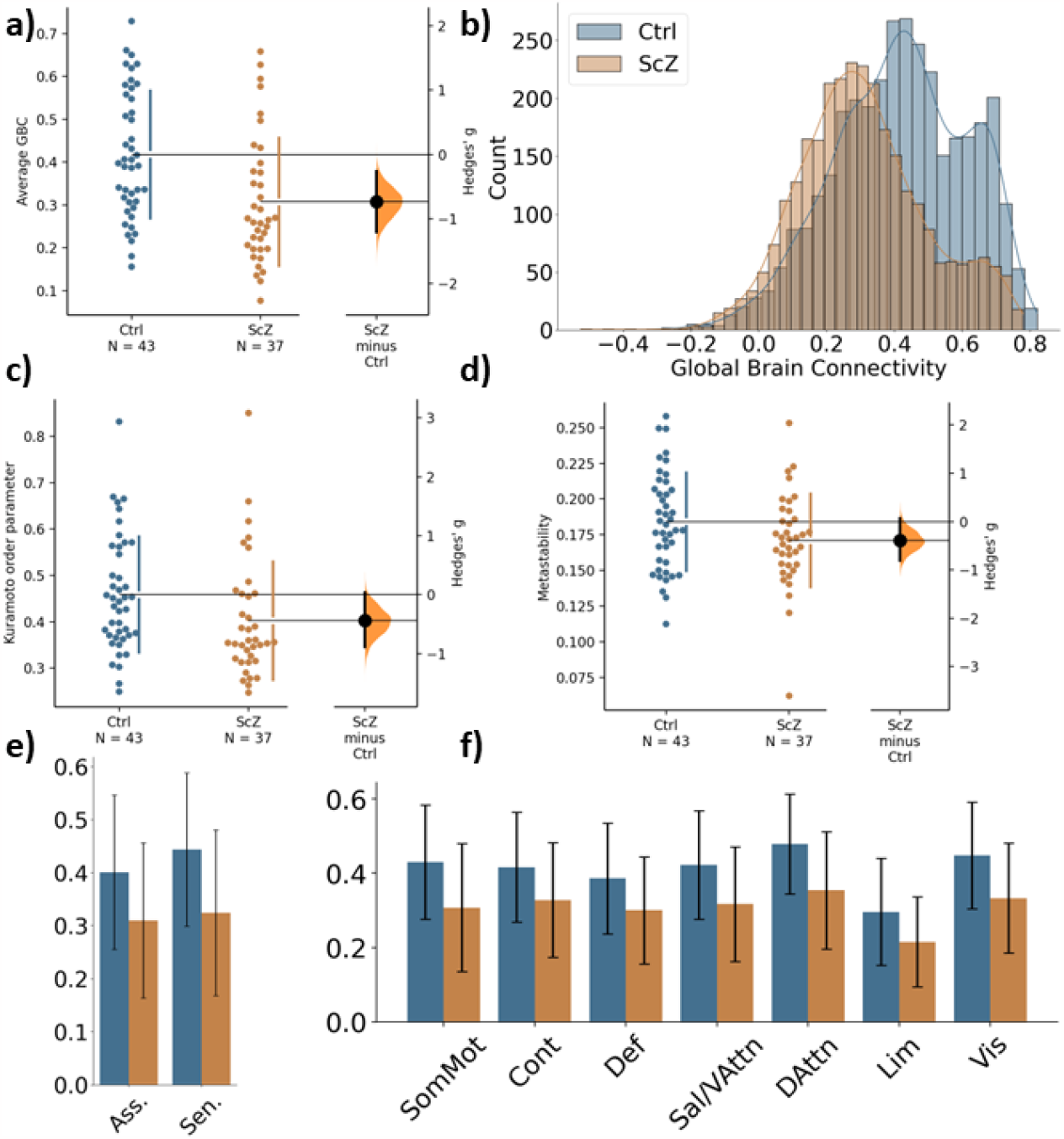
Global differences in functional connectivity and temporal dynamics between healthy controls and ScZ patients. a) Comparison of average GBC per participant for the two groups. Individual dots represent average GBC for one participant. The difference plot on the right shows the difference between the groups in terms of effect size. b) Histogram of region-wise GBC values for the two groups. The histogram displays the region-wise GBC data pooled for all participants in each group. c) Synchrony comparison between the two groups. Each dot represents the mean Kuramoto order parameter (a measure of synchrony) for one participant. The difference plot on the right shows the group difference in terms of effect size. d) Metastability comparison between the two groups. Each dot represents the metastability of one participant. The difference plot on the right shows the group difference in terms of effect size. e) Comparison of global brain connectivity for association areas (Asso. comprising: DMN, Cont, Sal/VAttn) and sensory areas (Sen. comprising: Sommot, Vis, DAttn). f) Comparison of global brain connectivity for the seven functional networks from Yeo et al. (2011): Somato-motor subnetwork (SomMot), Control subnetwork (Cont), Default mode subnetwork (Def), Salience/Ventral attention subnetwork (Sal/VAttn), Dorsal attention subnetwork (DAttn), Limbic subnetwork (Lim), Visual subnetwork (Vis).

**Figure 2.**
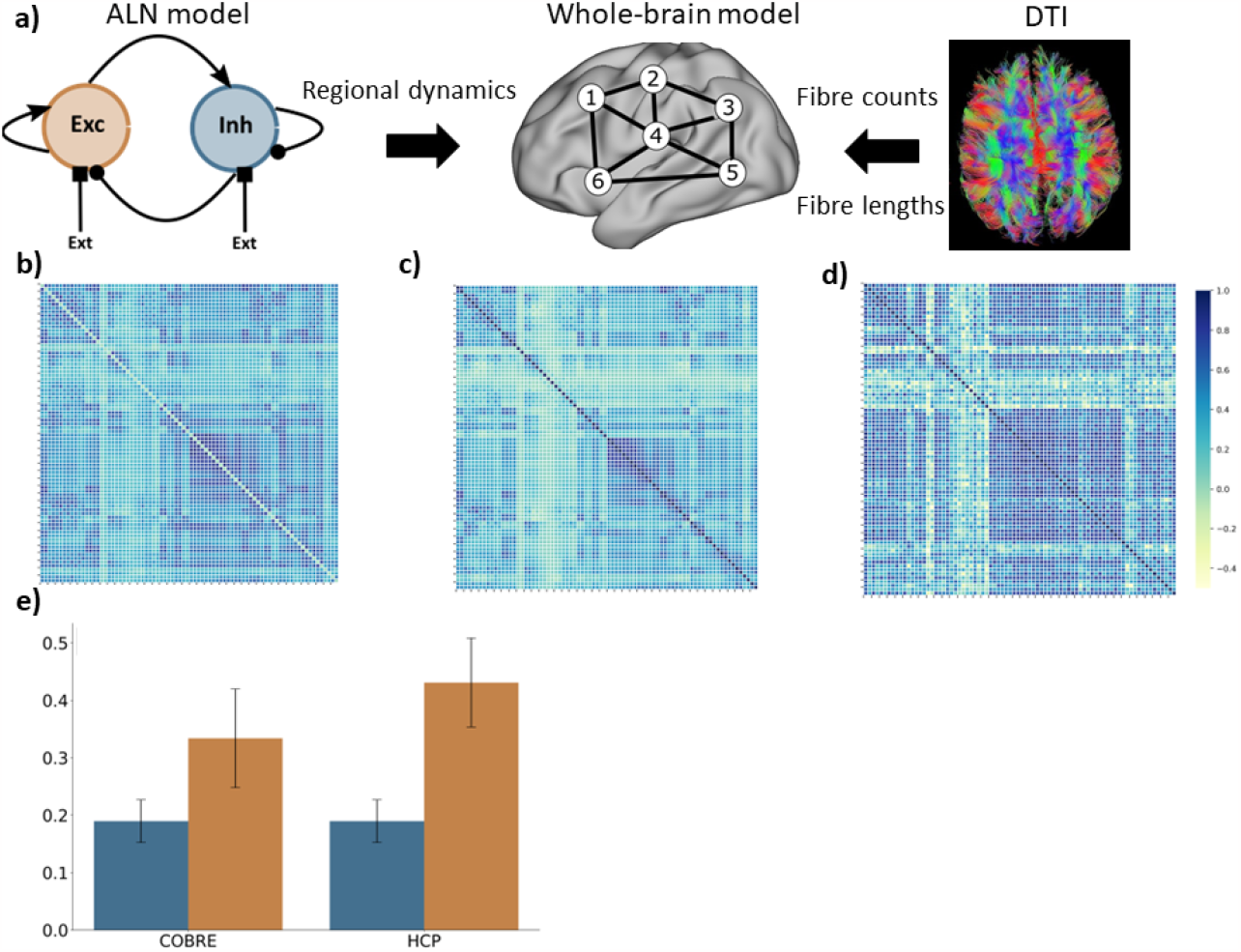
Computational model. a) Modelling approach combining a model for the regional dynamics with anatomical input that defines the structural network. b) Average FC matrix for the COBRE sample c) Average FC matrix for the HCP sample d) Model FC matrix e) comparison of the correlation of empSC to empFC (blue) and the correlation of simFC and empFC (yellow) for the COBRE (left) and the for the HCP (right) data sets.

#### 2.5.2 BOLD model

In order to compare the model output, i.e. the neural activity of the regions, to the BOLD signal of the rs-fMRI data, the firing rates of the excitatory population of each region had to be converted into model BOLD signal timecourses. Here, we used the well-established Balloon-Windkessel model (Friston et al. (2000); Deco et al. (2013)), for specific parameters see Friston et al. (2003).

#### 2.5.3 Network connectivity

Structural images were preprocessed employing a semi-automatic pipeline implemented in the FSL toolbox (www.fmrib.ox.ac.uk/fsl, FMRIB, Oxford). For the anatomical T1-weighted images we used the brain extraction toolbox (BET) in FSL to remove non-brain tissue and to generate the brain masks. After manual quality checks, 80 cortical regions were defined according to the automatic anatomical labelling (AAL2) atlas (Rolls et al. (2015)). For the diffusion-weighted images, we performed a brain extraction as well and corrected the images for head motion and eddy current distortions afterwards. Probabilistic fibre tracking, using the Bayesian Estimation of Diffusion Parameters Obtained using Sampling Techniques (BEDPOSTX) and the PROBTRACKX algorithms implemented in FSL (Behrens et al. (2007)), was then used with 5,000 random seeds per voxel to extract individual connectomes. Since the tractography does not yield directionality information and the connectome matrices are non-symmetric, we explicitly enforced symmetry by averaging the entries from region i to region j and from region j to region i for all pairs of regions. Furthermore, we normalised each connectome by dividing each matrix entry by the maximum matrix entry, thus ensuring compatibility between participants. The resulting connectome then determines the relative coupling strength between regions in the above described computational whole-brain model. The fibre tracking also yielded matrix fibre lengths for each participant, which, when multiplied with the signal speed, determines the delay of signal propagation between any two regions in the model.

#### 2.5.4 Modelling ScZ-associated changes

We implemented four different sets of parameter changes that are thought to represent the following four ScZ-associated alterations: 1) local GABAergic inhibition, 2) local glutamatergic excitation of inhibitory cells, 3) global interregional coupling, and 4) global noise levels.

First, we systematically reduced GABAergic inhibition in the model. Postmortem gene expression studies have robustly demonstrated reduced levels of parvalbumine (PV) and somatostatin (SST) expression in PV (Hashimoto et al. (2003)) and SST (Morris et al. (2008)) interneurons together with a reduction of GAD_65_ and GAD_67_ (Hashimoto et al. (2003)), in cortical regions in ScZ. We implemented these changes as a reduction of the inhibitory weights *J*_*EI*_ and *J*_*II*_ in the ALN model of the regional dynamics. We varied the strength of the inhibition onto the excitatory population *J*_*EI*_ and onto the inhibitory population *J*_*II*_ simultaneously in the range from 100% to 60% in steps of 5%.

Next, we systematically reduced the glutamatergic, excitatory drive onto inhibitory neurons in our model of regional activity. These changes reflected the reduced and more varied colocalization of glutamatergic pre- and postsynaptic markers on PV interneurons in dorsolateral prefrontal cortex (DLPFC) (Chung et al. (2016, 2022)). Specifically, we reduced the excitatory weight onto inhibitory neurons *J*_*IE*_ in the ALN model in a range from 100% to 60% in steps of 5%.

Global dysconnectivity might also be explained by a simple reduction of the global connectivity strength. Therefore, to test whether the differences we found experimentally could alternatively be explained by an overall network decoupling, we reduced the global coupling strength *K*_*gl*_ in the range from 100% to 60% in steps of 5%.

Finally, the global alterations of functional connectivity might also be the result of an increase in background noise disrupting functional connectivity in the network (Winterer et al. (2000); Winterer and Weinberger (2004); Winterer et al. (2004)). Consequently, we increased the global background noise *σ*_*ou*_ in a range from 100% to 140% in steps to 5%, to test whether a global increase in noise level can account for the connectivity differences found in the experimental data.

#### 2.5.5 Simulation details

Simulations were implemented using the neurolib Python framework (Cakan et al. (2021)). The differential equations of the model were solved numerically using an Euler forward scheme with a time step of 0.1 ms. For all described simulations the duration was 70 s and we discarded the transient response in the first 5 s before calculating any of the above described measures. To assess the robustness of our results, we created 40 virtual subjects by changing the seed for the random number generator underlying the Ornstein-Uhlenbeck noise process. These 40 virtual subjects were then kept fixed for all implemented changes allowing for a direct comparison to the default, ‘healthy’ condition.

## 3 RESULTS

### 3.1 Demographic and clinical characteristics

The control and the patient group did not differ significantly in terms of age and gender (see Table 1). Patients also did not show a change in symptomatology or type/dose of antipsychotic medication during the three months before the assessment (for more details see Aine et al. (2017)).

### 3.2 Global differences in connectivity strength and temporal dynamics between ScZ patients and healthy controls

Global GBC was significantly reduced in patients with ScZ (effect size *g* = −0.65; see Figure 1 a) and Table 3). Comparing both groups a substantial shift from high GBC towards medium to low GBC values occurs in ScZ patients (Figure 1 b) and Table 3). Synchrony, as measured by the Kuramoto order parameter was lower in the patient group (effect size *g* = −0.44; see Figures 1 c) and Table 3). However, variability in synchrony, measured by metastability, did not significantly differ between groups (Figure 1 d) and Table 3).

**Table 3.** Local and global group differences. Overview of the global and local differences in functional connectivity and temporal dynamics between the healthy control and the ScZ patient group.

|  | Mean difference | Hedges' g | 95% CI | p value |
| --- | --- | --- | --- | --- |
| Global cortical GBC | -0.11 | -0.65 | [-1.11 -0.18] | p=0.0056 |
| Global cortical synchrony | -0.12 | -0.44 | [-0.89 0.04] | p=0.0488 |
| Global cortical metastability | -0.005 | -0.39 | [-0.81 0.07] | p=0.0850 |
| GBC Sensory areas | -0.12 | -0.78 | [-1.26 -0.29] | p=0.0004 |
| GBC Association areas | -0.09 | -0.61 | [-1.07 -0.14] | p=0.0076 |
| GBC Somato-motor (SomMot) | -0.12 | -0.74 | [-1.20 -0.26] | p=0.0008 |
| GBC Control (Cont) | -0.09 | -0.57 | [-1.03 -0.10] | p=0.0110 |
| GBC Default mode (Def) | -0.09 | -0.57 | [-1.03 -0.11] | p=0.0110 |
| GBC Salience/Ventral attention (Sal/VAtn) | -0.11 | -0.69 | [-1.14 -0.21] | p=0.0024 |
| GBC Dorsal attention (DAtn) | -0.12 | -0.83 | [-1.31 -0.33] | p=0.0001 |
| GBC Limbic (Lim) | -0.08 | -0.59 | [-1.02 -0.13] | p=0.0102 |
| GBC Visual (Vis) | -0.11 | -0.77 | [-1.26 -0.29] | p=0.0010 |

Reductions of functional connectivity strengths affected all seven subnetworks (effect sizes ranging from *g* = −0.57 to *g* = −0.83; see Table 3), with the dorsal-attention, the somato-motor and the visual subnetworks showing the strongest effects (Figure 1 f)).

We further tested whether the GBC differences we found were specific to association areas as indicated by a previous study (Yang et al. (2016)). We grouped the default mode subnetwork, the control subnetwork and the ventral attention subnetwork together as the association areas and the somatomotor subnetwork, the visual subnetwork and the dorsal attention subnetwork as the sensory areas. We found reduced GBC for ScZ patients in both groupings, with the sensory areas showing an even stronger effect than the association areas (effect sizes *g* = −0.78 for sensory areas versus *g* = −0.61 for association areas, see Figure 1 e) and Table 3).

### 3.3 Mechanisms underlying connectivity and dynamics alterations

#### 3.3.1 Control model

We derived a model of healthy large-scale cortical activity that matched the behaviour of the control group data from the COBRE study well in terms of functional connectivity (Figures 2 b) and d)). The correlation between simulated FC (simFC) and empirical FC (empFC) (*r* = 0.33 ± 0.09; Figure 2 e)) was higher than the correlation between empirical structural (empSC) and empFC (*r* = 0.19 ± 0.07; Figure2 e)).

To further assert that the default model captures the resting-state functional connectivity of healthy subjects well, we also validated the model behaviour against a set of 156 subjects from the HCP S1200 release. Here, we also found a good fit for functional connectivity (Figure 2 c) and d)).

Overall, the model functional connectivity correlated well with the empirical functional connectivity of individual HCP subjects (*r* = 0.43 ± 0.08; see Figure 2 e)). This correlation was again substantially higher than the correlation of structural connectivity and empirical functional connectivity (*r* = 0.20 ± 0.08; see Figure 2 e)).

#### 3.3.2 Modelling ScZ-associated alterations

We systematically performed perturbations to four key aspects of the model that have been associated with schizophrenia: 1) local GABAergic inhibition, 2) local glutamatergic excitation of inhibitory cells, 3) global interregional coupling, and 4) global noise levels.

We found that changing the inhibitory weights (model perturbation 1) did not alter the global GBC and the GBCs for sensory and association areas significantly. Furthermore, the changes did not alter the synchrony and the metastability (see Supplementary Table S3). As for the local changes to the inhibitory system, changes to the glutamatergic excitatory drive to the inhibitory population (model perturbation 2) did not result in significant changes in GBC on all levels, synchrony and metastability (see Supplementary Table S4).

A reduction of global coupling (model perturbation 3) resulted in a strong decrease in global brain connectivity as well as connectivity within the sensory and association systems (Table 4). Additionally, synchrony decreased strongly and metastability increased for larger reductions (Table 4).

**Table 4.**
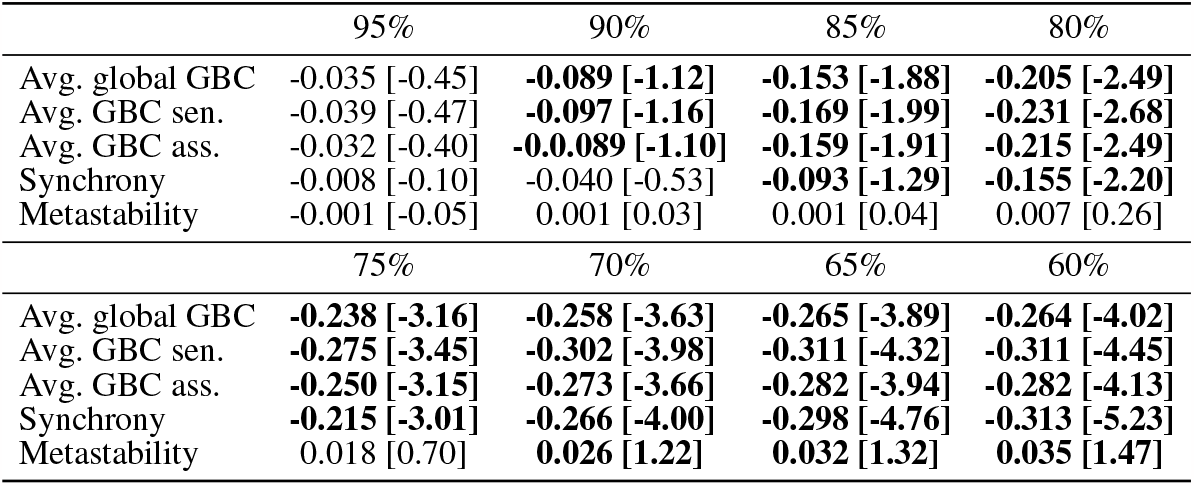
ScZ-associated changes of global coupling. Comparison of average global GBC, average GBC in sensory areas, average GBC in association areas, average synchrony and average metastability for reduced global coupling (from 95% to 60% in steps of 5%). Shown are the mean differences, i.e. the mean of the default condition minus the respective reduced global coupling condition and in brackets the effect size (Hedge’s g). The mean in each condition is calculated over the 40 virtual subjects. Significant differences, i.e. a permutation *p* value of *<* 0.001, are highlighted in bold. Permutation tests were performed using 5,000 permutations of labels.

|  | 95% | 90% | 85% | 80% |
| --- | --- | --- | --- | --- |
| Avg. global GBC | -0.035 [-0.45] | **-0.089 [-1.12]** | **-0.153 [-1.88]** | **-0.205 [-2.49]** |
| Avg. GBC sen. | -0.039 [-0.47] | **-0.097 [-1.16]** | **-0.169 [-1.99]** | **-0.231 [-2.68]** |
| Avg. GBC ass. | -0.032 [-0.40] | **-0.089 [-1.10]** | **-0.159 [-1.91]** | **-0.215 [-2.49]** |
| Synchrony | -0.008 [-0.10] | -0.040 [-0.53] | **-0.093 [-1.29]** | **-0.155 [-2.20]** |
| Metastability | -0.001 [-0.05] | 0.001 [0.03] | 0.001 [0.04] | 0.007 [0.26] |
|  | 75% | 70% | 65% | 60% |
| Avg. global GBC | **-0.238 [-3.16]** | **-0.258 [-3.63]** | **-0.265 [-3.89]** | **-0.264 [-4.02]** |
| Avg. GBC sen. | **-0.275 [-3.45]** | **-0.302 [-3.98]** | **-0.311 [-4.32]** | **-0.311 [-4.45]** |
| Avg. GBC ass. | **-0.250 [-3.15]** | **-0.273 [-3.66]** | **-0.282 [-3.94]** | **-0.282 [-4.13]** |
| Synchrony | **-0.215 [-3.01]** | **-0.266 [-4.00]** | **-0.298 [-4.76]** | **-0.313 [-5.23]** |
| Metastability | 0.018 [0.70] | **0.026 [1.22]** | **0.032 [1.32]** | **0.035 [1.47]** |

An increase in noise levels (model perturbation 4) yielded a strong decrease in global brain connectivity as well as connectivity within the sensory and association systems, even stronger than for the global coupling changes (Table 5). Additionally, synchrony decreased strongly and metastability increased for larger noise strengths (Table 5).

**Table 5.** ScZ-associated changes of noise parameters. Comparison of average global GBC, average GBC in sensory areas, average GBC in association areas, average synchrony, and average metastability with increased noise (from 105% to 140% in steps of 5%). Shown are the mean differences, i.e. the mean of the default condition minus the respective increased noise condition and in brackets the effect size (Hedge’s g). The mean in each condition is calculated over the 40 virtual subjects. Significant differences, i.e. a permutation *p* value of *<* 0.001, are highlighted in bold. Permutation tests were performed using 5,000 permutations of labels.

|  | 105% | 110% | 115% | 120% |
| --- | --- | --- | --- | --- |
| Avg. global GBC | **-0.078 [-0.98]** | **-0.129 [-1.60]** | **-0.199 [-2.56]** | **-0.260 [-3.56]** |
| Avg. GBC sen. | **-0.082 [-0.98]** | **-0.139 [-1.64]** | **-0.215 [-2.59]** | **-0.285 [-3.64]** |
| Avg. GBC ass. | **-0.078 [-0.95]** | **-0.133 [-1.58]** | **-0.204 [-2.51]** | **-0.268 [-3.57]** |
| Synchrony | -0.050 [-0.67] | **-0.080 [-1.05]** | **-0.121 [-1.83]** | **-0.168 [-2.42]** |
| Metastability | 0.008 [0.26] | 0.006 [0.21] | 0.010 [0.37] | 0.014 [0.52] |
|  | 125% | 130% | 135% | 140% |
| Avg. global GBC | **-0.313 [-4.58]** | **-0.362 [-5.71]** | **-0.383 [-6.07]** | **-0.394 [-6.08]** |
| Avg. GBC sen. | **-0.345 [-4.86]** | **-0.399 [-6.01]** | **-0.424 [-6.39]** | **-0.433 [-6.30]** |
| Avg. GBC ass. | **-0.323 [-4.56]** | **-0.373 [-5.62]** | **-0.396 [-6.15]** | **-0.408 [-6.22]** |
| Synchrony | **-0.202 [-2.97]** | **-0.230 [-3.62]** | **-0.244 [-3.95]** | **-0.261 [-4.17]** |
| Metastability | 0.018 [0.70] | **0.019 [0.75]** | **0.026 [1.03]** | **0.027 [1.02]** |

## 4 DISCUSSION

### 4.0.1 Global changes in connectivity and temporal dynamics

Evidence for large-scale dysconnectivity in functional networks has been accumulated over the last years in ScZ (Liang et al. (2006); Bluhm et al. (2007); Bassett et al. (2008); Liu et al. (2008); Bullmore and Sporns (2009)). However, it is still unclear, how these changes relate to changes on the microscopic level. To address this gap, we analysed resting-state fMRI data from healthy participants and patients with chronic ScZ. We identified a global reduction in functional connectivity that affected both sensory and association areas equally and that was present for all functional subnetworks together with a moderate decrease of temporal synchrony. Using a biophysical network model, we found that a decrease in global coupling or an increase in global noise levels could explain the connectivity reduction and the increase in synchrony best, whereas local changes to the glutamatergic or GABAergic system did not produce changes matching our experimental findings. However, both changes also yielded an increase in metastability in our model, which we did not find in the experimental data.

Our findings of reduced global brain connectivity are in line with previous research. For example, Lynall et al. (2010) and Bassett et al. (2012) both found significantly reduced global integration in patients with schizophrenia. However, we did not find stronger connectivity disturbances in association areas compared to sensory areas, as previously reported (Yang et al. (2016)).

Our analysis of the temporal dynamics of the activity, i.e. synchrony and metastability, revealed a decrease in synchrony but no change in metastability. Our finding of unchanged metastability is in line with previous findings of Lee et al. (2018) on the same dataset but in contrast to very recent work from Hancock et al. (2023a), proposing metastability as a candidate biomarker for schizophrenia. However, we have to note that Hancock et al. (2023a) introduced a new measure of metastability with increased sensitivity to detect the differences between healthy controls and ScZ patients. This new measure of metastability did not rely on predefined brain parcellations but rather flexibly defined recurring spatio-temporal modes, so-called ‘communities’ where single brain regions may be grouped into more than one community. As this approach was not applicable to our computational network model we did not employ it in our analysis. Overall, several different metastability measures have been proposed and have been applied in different contexts in neuroscience (Hancock et al. (2023b)).

### 4.0.2 Mechanistic explanations of global changes in ScZ

Reduced global coupling and increased global noise levels are in line with earlier modelling studies. For example, several studies, using both simple phase oscillator models and dynamic mean-field models, have shown that a decrease of global coupling compared to the best model fit to human resting-state data led to a decrease in connectivity and a more random, less integrated graph structure (Cabral et al. (2012a, 2013, 2012b)). Similar to the model presented here, the operating point is chosen close to a bifurcation point from a silent down state to a limit-cycle which produces oscillating activity. In this regime, both functional connectivity and temporal dynamics best match empirical data. Therefore, the reduced coupling or the increased global noise disturbs this specific state and thus reduces global connectivity, synchrony and more complex network properties.

Previous work on the effects of changes to the glutamatergic and GABAergic system has demonstrated profound alterations on the cortical microcircuit level. For example, numerous computational studies have shown that ScZ-associated changes on the microcircuit level can lead to substantial reductions in gamma power in auditory steady-state response tasks (Metzner et al. (2016); Metzner and Steuber (2021); Metzner et al. (2019); Vierling-Claassen et al. (2008)). Since local gamma oscillations have been hypothesized to at least partially determine the large-scale functional connectivity and temporal dynamics of resting-state activity (Cabral et al. (2014, 2022)), it seems surprising that changes to either of the systems did not produce changes in global brain connectivity in our model. One reason for the lack of impact of the changes might be that we applied them homogeneously. In the work presented here, we only varied glutamatergic or GABAergic strength globally, i.e. without any spatial heterogeneity. Therefore, it seems plausible that these changes disturbed the local, regional nodes all in a similar fashion and thereby did not substantially alter their interrelation, thus not changing global brain connectivity. Indeed, several studies have demonstrated that heterogeneous models of cortex, which explicitly incorporate regional differences in dynamics, match experimental resting-state functional connectivity more accurately (Demirtaş et al. (2019); Kong et al. (2021)). Importantly, these regional differences in dynamics covary with expression profiles for markers of glutamatergic and GABAergic neurotransmission and E-I balance (Burt et al. (2018); Demirtaş et al. (2019)). Therefore, a more detailed, heterogeneous model might be able to shed more light on the effect of E-I balance changes associated with ScZ on large-scale functional networks.

### 4.0.3 Limitations

The computational model that we have employed in this study, while generally showing a very good fit to the experimental data, is not fully biophysically realistic. Moreover, the model used an average connectome and was not able to provide subject-specific, individual results for each participant. Furthermore, the anatomic parcellation (AAL2 Rolls et al. (2015)) is relatively coarse-grained with a number of 80 cortical regions.

The ALN model that was used to simulate regional activity has been demonstrated to approximate cortical resting-state activity (Cakan and Obermayer (2020); Cakan et al. (2022)). However, it is restricted to the cortex. Including subcortical regions such as the thalamus into whole-brain models is still in its infancy and rarely goes beyond coupling a single cortical and thalamic region (e.g. Jajcay et al. (2022), but see Griffiths et al. (2020)).

The ALN model also presents a simplification of the regional circuitry as it approximates and neglects both the variability of cell types, especially the diversity of inhibitory interneurons, and the laminar structure of the cortex. Therefore, the inclusion of more detailed models of regional activity, both in terms of cell type diversity and of laminar structure and connectivity, seems likely to further our understanding of ScZ dysconnectivity and its underlying mechanisms.

Lastly, the regional ALN model we used had the same parameters regardless of the cortical region it represented, i.e. we implemented a homogeneous model in that respect. As already discussed above, cortical regions are known to differ in various important aspects, whose incorporation are likely to provide additional insight into the pathophysiology of schizophrenia.

#### 4.0.4 Conclusion

The current study provides further evidence of large-scale changes in connectivity and temporal dynamics in ScZ through the analysis of resting-state fMRI. Furthermore, through computational modelling, it provides novel evidence that these changes are likely the result of global reductions in coupling or increases noise levels and not of changes to local recurrent connectivity. These findings emphasize the effect of global alterations in ScZ and have possible implications for the development of treatments.

## Data Availability

All data produced are available online at https://github.com/ChristophMetzner/FrontiersPsychiatry2023

## DATA AVAILABILITY STATEMENT

The datasets analyzed for this study can be obtained via *schizconnect.org*. The code for the computational model can be found here *https://github.com/ChristophMetzner/FrontiersPsychiatry2023*.

## 7 SUPPLEMENTARY TABLES AND FIGURES

### 7.1 Figures

**Figure 1.**
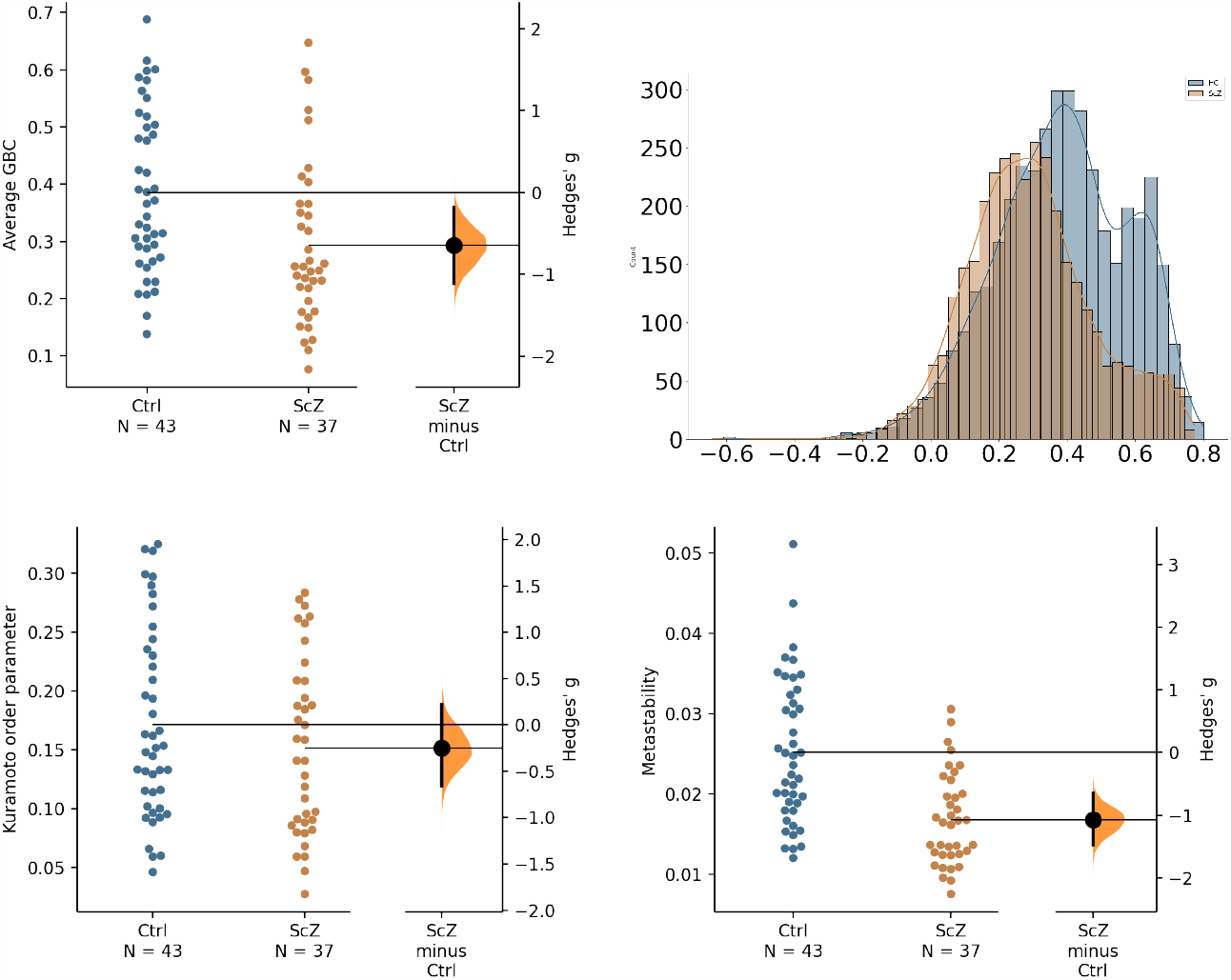
Global differences in functional connectivity and temporal dynamics. a) Comparison of average GBC per participant for the two groups. Individual dots represent average GBC for one participant. The difference plot on the right shows the difference between the groups in terms of effect size. b) Histogram of region-wise GBC values for the two groups. The histogram displays the region-wise GBC data pooled for all participants in each group. c) Synchrony comparison between the two groups. Each dot represents the mean Kuramoto order parameter (a measure of synchrony) for one participant. The difference plot on the right shows the group difference in terms of effect size. d) Metastability comparison between the two groups. Each dot represents the metastability of one participant. The difference plot on the right shows the group difference in terms of effect size.

### 7.2 Tables

**Table 6.** Regional effect sizes. Effect sizes and p values for the comparison of the control and the patient group in average GBC for each of the 90 regions of the AAL2 parcellation.

| Region | Hedges' g | p value |
| --- | --- | --- |
| Precentral L | -0.69 | 0.0024 |
| Precentral R | -0.68 | 0.0024 |
| Frontal Sup 2 L | -0.70 | 0.0018 |
| Frontal Sup 2 R | -0.49 | 0.0318 |
| Frontal Mid 2 L | -0.56 | 0.0126 |
| Frontal Mid 2 R | -0.43 | 0.0608 |
| Frontal Inf Oper L | -0.72 | 0.0014 |
| Frontal Inf Oper R | -0.67 | 0.0042 |
| Frontal Inf Tri L | -0.68 | 0.0030 |
| Frontal Inf Tri R | -0.59 | 0.0092 |
| Frontal Inf Orb 2 L | -0.53 | 0.0212 |
| Frontal Inf Orb 2 R | -0.41 | 0.0644 |
| Rolandic Oper L | -0.61 | 0.0078 |
| Rolandic Oper R | -0.67 | 0.0032 |
| Supp Motor Area L | -0.75 | 0.0006 |
| Supp Motor Area R | -0.84 | 0.0006 |
| Olfactory L | -0.26 | 0.2536 |
| Olfactory R | -0.16 | 0.4704 |
| Frontal Sup Medial L | -0.51 | 0.0242 |
| Frontal Sup Medial R | -0.52 | 0.0206 |
| Frontal Med Orb L | -0.40 | 0.0736 |
| Frontal Med Orb R | -0.42 | 0.0600 |
| Rectus L | -0.52 | 0.0264 |
| Rectus R | -0.32 | 0.1518 |
| OFCmed L | -0.47 | 0.0404 |
| OFCmed R | -0.31 | 0.1728 |
| OFCant L | -0.30 | 0.1932 |
| OFCant R | -0.28 | 0.2026 |
| OFCpost L | -0.43 | 0.0578 |
| OFCpost R | -0.51 | 0.0186 |
| OFClat L | -0.60 | 0.0094 |
| OFClat R | -0.13 | 0.5442 |
| Insula L | -0.74 | 0.0018 |
| Insula R | -0.65 | 0.0050 |
| Cingulate Ant L | -0.49 | 0.0296 |
| Cingulate Ant R | -0.51 | 0.0248 |
| Cingulate Mid L | -0.87 | 0.0002 |
| Cingulate Mid R | -0.82 | 0.0002 |
| Cingulate Post L | -0.57 | 0.0132 |
| Cingulate Post R | -0.35 | 0.1174 |
| Calcarine L | -0.82 | 0.0002 |
| Calcarine R | -0.76 | 0.0016 |
| Cuneus L | -0.77 | 0.0010 |
| Cuneus R | -0.61 | 0.0064 |
| Lingual L | -0.89 | <0.0001 |
| Lingual R | -0.85 | 0.0002 |

**Table 7.** Regional effect sizes - ctd. Effect sizes and p values for the comparison of the control and the patient group in average GBC for each of the 90 regions of the AAL2 parcellation.

| Region | Hedges' g | p value |
| --- | --- | --- |
| Occipital Sup L | -0.71 | 0.0012 |
| Occipital Sup R | -0.72 | 0.0024 |
| Occipital Mid L | -0.85 | 0.0002 |
| Occipital Mid R | -0.92 | <0.0001 |
| Occipital Inf L | -0.84 | 0.0004 |
| Occipital Inf R | -0.56 | 0.014 |
| Fusiform L | -0.89 | 0.0002 |
| Fusiform R | -0.98 | <0.0001 |
| Postcentral L | -0.70 | 0.0024 |
| Postcentral R | -0.70 | 0.002 |
| Parietal Sup L | -0.85 | <0.0001 |
| Parietal Sup R | -0.94 | <0.0001 |
| Parietal Inf L | -0.85 | <0.0001 |
| Parietal Inf R | -0.51 | 0.025 |
| SupraMarginal L | -0.56 | 0.0132 |
| SupraMarginal R | -0.61 | 0.0072 |
| Angular L | -0.53 | 0.0184 |
| Angular R | -0.51 | 0.0234 |
| Precuneus L | -0.88 | 0.003 |
| Precuneus R | -0.69 | 0.001 |
| Paracentral Lobule L | -0.72 | 0.0126 |
| Paracentral Lobule R | -0.58 | 0.0106 |
| Heschl L | 0.59 | 0.0028 |
| Heschl R | -0.70 | <0.0001 |
| Temporal Sup L | -0.96 | <0.0001 |
| Temporal Sup R | -0.83 | 0.0076 |
| Temporal Pole Sup L | -0.6 | 0.001 |
| Temporal Pole Sup R | -0.77 | 0.001 |
| Temporal Mid L | -0.81 | 0.0008 |
| Temporal Mid R | -0.87 | <0.0001 |
| Temporal Pole Mid L | -0.68 | 0.0026 |
| Temporal Pole Mid R | -0.77 | 0.0008 |
| Temporal Inf L | -0.37 | 0.0972 |
| Temporal Inf R | -0.6 | 0.009 |

**Table 8.** ScZ-associated changes of GABA parameters. Comparison of average global GBC, average GBC in sensory areas, average GBC in association areas, average synchrony and average metastability for different conditions with reduced GABAergic output from 95% to 60% in steps of 5%. Shown are the mean differences (i.e. the mean of the default condition minus the respective reduced GABA condition. The mean in each condition is calculated over the 40 virtual subjects.) and in brackets the effect size (Hedge’s g). Significant differences, i.e. a permutation *p* value of *<* 0.001 is highlighted in bold. Permutation tests were performed using 5,000 permutations of labels.

|  | 95% | 90% | 85% | 80% |
| --- | --- | --- | --- | --- |
| Avg. global GBC | 0.001 [0.02] | 0.003 [0.04] | 0.003 [0.04] | 0.003 [0.04] |
| Avg. GBC sen. | 0.002 [0.02] | 0.003 [0.04] | 0.003 [0.03] | 0.002 [0.02] |
| Avg. GBC ass. | 0.001 [0.02] | 0.004 [0.04] | 0.005 [0.06] | 0.006 [0.06] |
| Synchrony | 0.003 [0.04] | 0.006 [0.08] | 0.008 [0.10] | 0.006 [0.08] |
| Metastability | -0.001 [-0.04] | -0.002 [-0.05] | -0.002 [-0.05] | -0.001 [-0.02] |
|  | 75% | 70% | 65% | 60% |
| Avg. global GBC | 0.004 [0.05] | 0.007 [0.08] | 0.005 [0.06] | 0.013 [0.16] |
| Avg. GBC sen. | 0.003 [0.03] | 0.005 [0.06] | 0.003 [0.03] | 0.012 [0.13] |
| Avg. GBC ass. | 0.007 [0.08] | 0.010 [0.11] | 0.008 [0.09] | 0.016 [0.19] |
| Synchrony | 0.009 [0.12] | 0.001 [0.01] | -0.003 [-0.04] | -0.005 [-0.06] |
| Metastability | -0.005 [-0.14] | -0.001 [-0.02] | 0.004 [0.13] | 0.007 [0.22] |

**Table 9.**
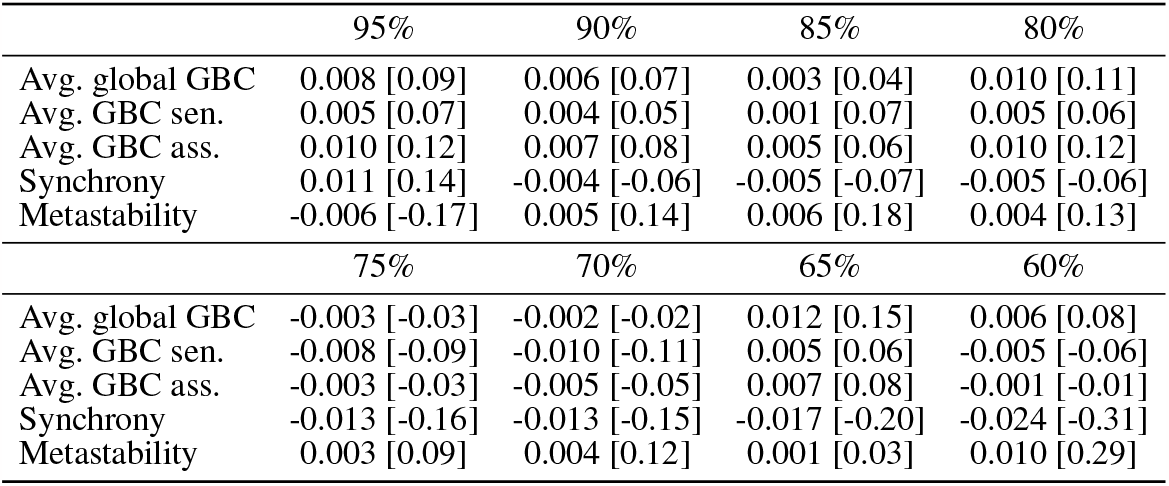
ScZ-associated changes of glutamatergic parameters. Comparison of average global GBC, average GBC in sensory areas, average GBC in association areas, average synchrony and average metastability for different conditions with reduced glutamatergic output from 95% to 60% in steps of 5%. Shown are the mean differences (i.e. the mean of the default condition minus the respective reduced glutamate condition. The mean in each condition is calculated over the 40 virtual subjects.) and in brackets the effect size (Hedge’s g). Significant differences, i.e. a permutation *p* value of *<* 0.001 is highlighted in bold. Permutation tests were performed using 5,000 permutations of labels.

|  | 95% | 90% | 85% | 80% |
| --- | --- | --- | --- | --- |
| Avg. global GBC | 0.008 [0.09] | 0.006 [0.07] | 0.003 [0.04] | 0.010 [0.11] |
| Avg. GBC sen. | 0.005 [0.07] | 0.004 [0.05] | 0.001 [0.07] | 0.005 [0.06] |
| Avg. GBC ass. | 0.010 [0.12] | 0.007 [0.08] | 0.005 [0.06] | 0.010 [0.12] |
| Synchrony | 0.011 [0.14] | -0.004 [-0.06] | -0.005 [-0.07] | -0.005 [-0.06] |
| Metastability | -0.006 [-0.17] | 0.005 [0.14] | 0.006 [0.18] | 0.004 [0.13] |
|  | 75% | 70% | 65% | 60% |
| Avg. global GBC | -0.003 [-0.03] | -0.002 [-0.02] | 0.012 [0.15] | 0.006 [0.08] |
| Avg. GBC sen. | -0.008 [-0.09] | -0.010 [-0.11] | 0.005 [0.06] | -0.005 [-0.06] |
| Avg. GBC ass. | -0.003 [-0.03] | -0.005 [-0.05] | 0.007 [0.08] | -0.001 [-0.01] |
| Synchrony | -0.013 [-0.16] | -0.013 [-0.15] | -0.017 [-0.20] | -0.024 [-0.31] |
| Metastability | 0.003 [0.09] | 0.004 [0.12] | 0.001 [0.03] | 0.010 [0.29] |

